# Feasibility and lessons learned on remote trial implementation from TestBoston, a fully remote, longitudinal, large-scale COVID-19 surveillance study

**DOI:** 10.1101/2021.10.28.21265624

**Authors:** Sarah Naz-McLean, Andy Kim, Andrew Zimmer, Hannah Laibinis, Jen Lapan, Paul Tyman, Jessica Hung, Christina Kelly, Himaja Nagireddy, Surya Narayanan-Pandit, Margaret McCarthy, Saee Ratnaparkhi, Henry Rutherford, Rajesh Patel, Scott Dryden-Peterson, Deborah T. Hung, Ann E. Woolley, Lisa A. Cosimi

## Abstract

**Importance:** Remote clinical trials may reduce barriers to research engagement resulting in more representative samples. A critical evaluation of this approach is imperative to optimize this paradigm shift in research.

**Objective:** To assess design and implementation factors required to maximize enrollment and retention in a fully remote, longitudinal COVID-19 testing study.

**Design:** Fully remote longitudinal study launched in October 2020 and ongoing; Study data reported through July 2021.

**Setting:** Brigham and Women’s Hospital, Boston MA

**Participants:** Adults, 18 years or older, within 45 miles of Boston, MA.

**Intervention:** Monthly and “on-demand” at-home SARS-CoV-2 RT-PCR and antibody testing using nasal swab and dried blood spot self-collection kits and electronic surveys to assess symptoms and risk factors for COVID-19.

**Main Outcomes:** Enrollment, retention, and lessons learned.

**Results:** Between October 2020 and January 2021, we enrolled 10,289 participants reflective of Massachusetts census data. Mean age was 47 years (range 18-93), 5855 (56.9%) were assigned female sex at birth, 7181(69.8%) reported being White non-Hispanic, 952 (9.3%) Hispanic/Latinx, 925 (9.0%) Black, 889 (8.6%) Asian, and 342 (3.3%) other and/or more than one race. Lower initial enrollment among Black and Hispanic/Latinx individuals required an adaptive approach, leveraging connections to the medical system, coupled with community partnerships to ensure a representative cohort. Longitudinal retention was higher among participants who were White non-Hispanic, older, working remotely, and with lower socioeconomic vulnerability. Considerable infrastructure, including a dedicated participant support team and robust technology platforms was required to reduce barriers to enrollment, promote retention, ensure scientific rigor, improve data quality, and enable an adaptive study design to increase real-world accessibility.

**Conclusions:** The decentralization of clinical trials through remote models offers tremendous potential to engage representative cohorts, scale biomedical research, and promote accessibility by reducing barriers common in traditional trial design. Our model highlights the critical role that hospital-community partnerships play in remote recruitment, and the work still needed to ensure representative enrollment. Barriers and burdens within remote trials may be experienced disproportionately across demographic groups. To maximize engagement and retention, researchers should prioritize intensive participant support, investment in technologic infrastructure and an adaptive approach to maximize engagement and retention.

**Trial Registration:** N/A

**Key Points:** *Question:* Longitudinal clinical studies typically rely on in-person interactions to support recruitment, retention, and implementation. We define factors that promote demographically representative recruitment and retention through implementation of a fully remote COVID-19 study.

*Findings:* Remote trial models can reduce barriers to research participation and engage representative cohorts. Recruitment was strengthened by leveraging the medical system. Implementation highlighted participant burdens unique to this model, underscoring the need for a significant participant support team, robust technological infrastructure, and an adaptive, iterative approach.

*Meaning:* As remote trials become more common following the COVID-19 pandemic, methodologies to ensure accessibility, representation, and efficiency are crucial.

## INTRODUCTION

Since its emergence in December 2019, the COVID-19 pandemic has struck a massive blow to world health and economic systems and exposed long-standing healthcare disparities with over 242 million confirmed infections and over 4.9 million deaths worldwide (1). Accessibility and uptake of testing has varied between geographic locations and socio-economic groups, with many communities with the highest rates of COVID-19 simultaneously experiencing the lowest rates of testing (2,3). In October 2020, TestBoston, a longitudinal COVID-19 at-home testing study, was launched to understand the prevalence and risk factors for infection in the greater Boston area by providing access to SARS-CoV-2 viral and antibody testing with linkage to medical care and contact tracing. We hypothesized a fully remote model could reach a larger number of participants, while improving access to COVID-19 testing and biomedical research for underserved communities.

Disparities in clinical trial enrollment, particularly among Black and Hispanic/Latinx communities, are well-documented (4–7). Barriers to participation range from structural factors including required time commitments, distance and transportation to clinical sites, language barriers, and hidden costs, to a legacy of fear and mistrust stemming from historical atrocities in biomedical research (6–13). Remote models may provide greater efficiency, increased scale, wider geographic catchment areas, and the ability to reach a more representative population, including those unable or unwilling to travel for in-person study visits (5,14–22).

In the United States, prior to the SARS-CoV-2 pandemic, hybrid models of research have practiced elements of remote data collection; however, fully remote, decentralized studies with remote enrollment and collection of biomedical samples are new to the landscape (23–25). During the pandemic, clinical trials have faced unprecedented logistical barriers including social distancing protocols, restructuring of clinical sites to accommodate inpatient surges, participants’ fear of potential exposure during study visits, reduction of in-person research staff, and policies deeming study visits non-essential, necessitating adoption of remote methods to sustain research (5,18,26–30). Rather than being constrained by these limitations, researchers have capitalized on the need to transform the landscape toward a more equitable and efficient future through implementation of remote study models (14,17,18,21,31,32). However, to date, there is minimal experience in defining best practices in this domain.

Here we present the methods used in launching and implementing a large-scale fully remote longitudinal at-home COVID-19 surveillance study. We highlight key successes, challenges, and critical lessons learned applicable to remote trial implementation regardless of disease domain.

## METHODS

### Study Design

Study eligibility included adults 18 years of age or older residing within a 45-mile radius of Boston, Massachusetts. Participants were recruited from the general public and through Mass General Brigham (MGB, formerly Partners Healthcare), a not-for-profit, integrated healthcare system with 14 affiliated hospitals via 1) “Physician invitation” for individuals who had seen any MGB physician within the previous 2 years, and included an introductory letter from the patient’s primary care or specialty provider; 2) “Direct invitation” for MGB patients who previously opted in to be contacted about research opportunities; or 3) “Volunteer invitation” for individuals who signed up through an MGB-wide research recruitment website listing studies available to the general public. Eligible individuals received an invitation letter containing a one-time code and instructions to visit the online study portal, enter the code, create an account, read and sign an online informed consent, input their mailing address, and respond to a brief demographic survey (33). Completion of this process triggered a kit to be automatically sent to the participant. Interested individuals were encouraged to contact study staff to request assistance with the consent process, or to enroll by phone if unable to access the online portal. Study materials were translated into nine languages spoken in the geographical catchment area.

Extensive community outreach included consultation with state and local health departments to align study priorities, education sessions with MGB primary care and specialty clinics, outreach to community-based clinics and testing sites, press-releases, local news and radio segments, and partnering with local places of worship and community leaders to deliver general information about COVID-19 and answer questions from the community.

At enrollment, participants had the option to receive a one-time test kit, or to join the longitudinal cohort and receive monthly test kits for six months. Participants could request an additional “on-demand” kit at any time following exposure to COVID-19 or development of COVID-like symptoms.

Test kits were assembled and shipped from GBF Inc, High Point, NC and included an anterior nasal swab for reverse transcriptase – polymerase chain reaction (RT-PCR) testing and dried blood spot supplies for detection of SARS-CoV-2 antibody (Figure 1). When completing a kit, participants were directed to log in to their online portal to report recent exposures, new results of COVID-19 testing outside of the study, presence of symptoms concerning for COVID-19, and COVID-19 vaccination status (Figure 2).

**Figure 1:**
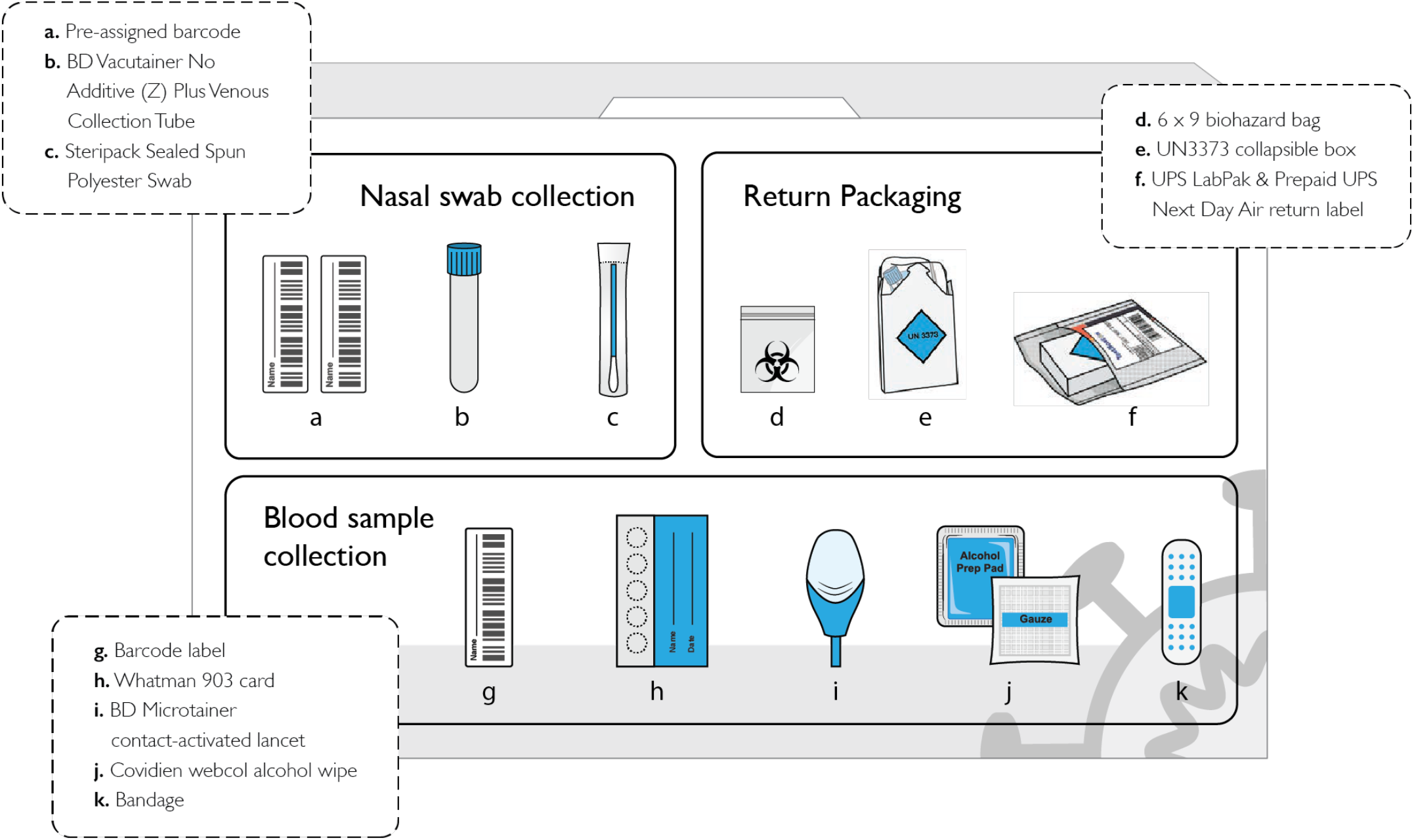
Schematic of TestBoston kit.

**Figure 2:**
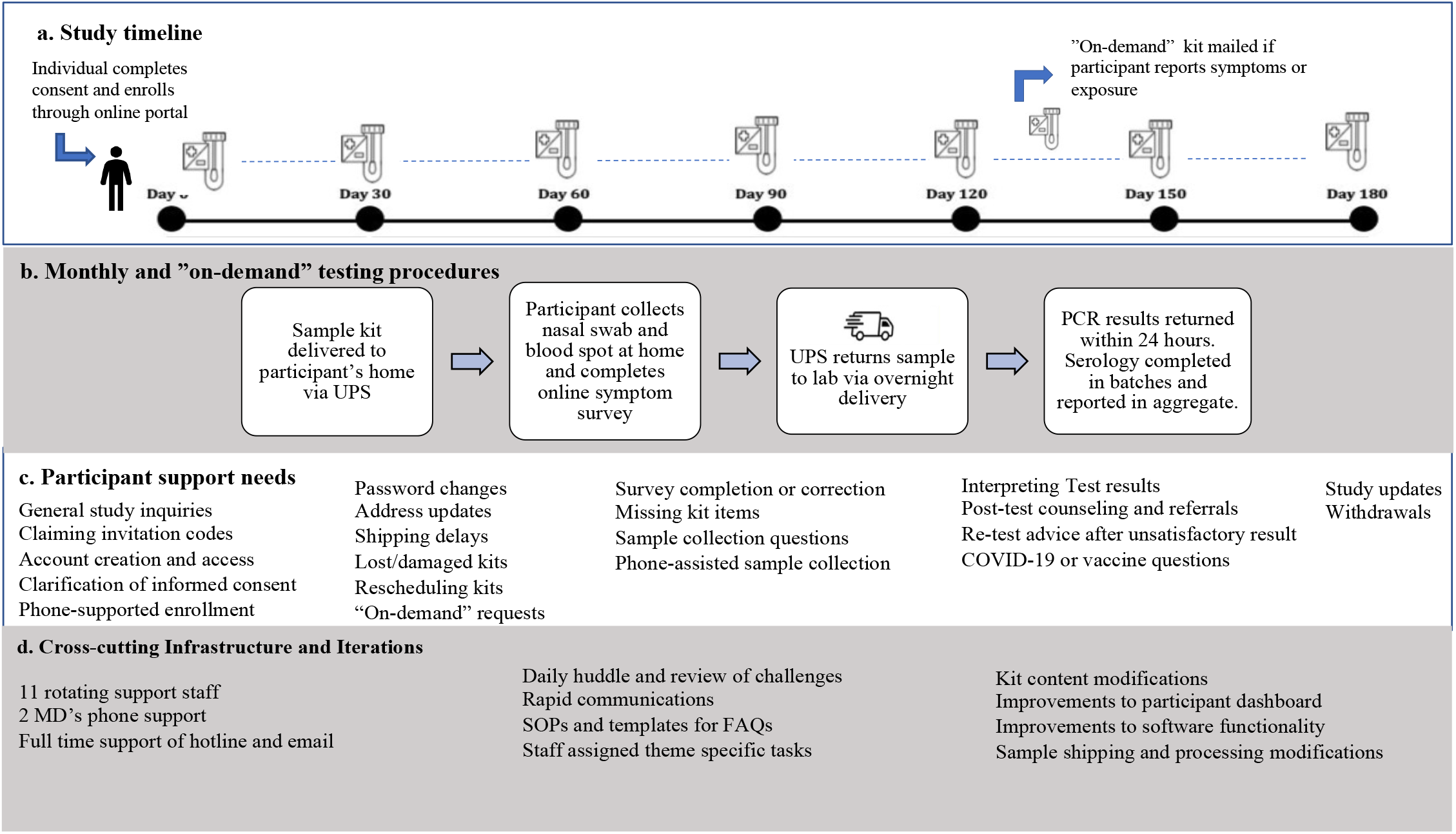
TestBoston procedures & related participant support requirements.

Participants returned completed kits to the lab overnight via a United Parcel Service (UPS) drop box or free home pickup. Upon arrival, kits were unboxed, reviewed for errors, then routed for high throughput RT-PCR testing in the COVID-19 Testing Program of Broad’s Clinical Research Sequencing Platform (CRSP). Viral RT-PCR results were delivered to participants’ online portals within 12-24 hours of sample receipt. Study staff notified individuals without computer access of their results by phone. Antibody results were reported in aggregate by zip code on the publicly available study website.

All positive RT-PCR results were reported directly to the Massachusetts Department of Public Health through the Massachusetts Epidemiologic Virtual Network (MAVEN), which triggered community contact tracing. A study physician contacted participants with positive results to offer post-test counseling and linkage to medical care through their primary care physician. Immediate referrals to either the emergency department or outpatient treatment of SARS-CoV-2, including monoclonal antibody treatments, were made based on current MGB guidelines (34).

### Data Systems

Online enrollment, consent, longitudinal data collection, kit shipping, tracking and receiving, and return of RT-PCR results were supported by Pepper, an open-source software product built on the Google Cloud Platform and maintained by the Broad Institute Data Sciences Platform to configure and operate direct-to-participant studies (35). Pepper provides Application Programming Interfaces (APIs) and user interfaces for participants, study team and logistical partners, utilizes 3rd party services, such as Auth0 for user authentication and authorization, SendGrid to distribute email communications to participants, and abides by all HIPAA security and breach rules. Participants were given a simple password-protected dashboard to complete study forms, request test kits, and view results. Data from Pepper was imported and supplemented with data from MGB medical records and stored in REDCap (Research Electronic Data Capture), a secure, web-based software platform designed to support data capture for research studies, hosted by MGB (36,37).

### Data Analysis

Longitudinal retention and engagement were measured based on number of kits returned, out of six, and time to kit return following delivery. We defined high level engagement as having completed five or more kits within 30 days of receipt; moderate engagement as 3 or 4 kits completed at any time point or 5 or more kits completed more than 30 days from receipt; and low engagement as having completed one or two kits at any time point. Multivariable Poisson regression was used with robust sandwich estimators to assess impact of the following baseline characteristics on level of engagement: sex, age (per 10 year increase), race and ethnicity, employment status (unemployed, employed remotely, or employed outside of home), and socioeconomic vulnerability as assessed by the Area Deprivation Index at census block group level (38). Statistical analysis was performed in R, version 4.1.1 (R Foundation).

### Ethical considerations

This study was approved by the MGB Institutional Review Board.

## RESULTS

### A multi-pronged approach leveraging links to the medical system enabled rapid, representative enrollment in absolute numbers, but exposed disparities in relative engagement

Between October 1, 2020 and February 2021, 102,576 people were invited to join the study, of which, 10,289 (10.0%) enrolled and returned at least one of six possible kits. “Direct invitations” were sent to 12,758 individuals who previously opted-in to be contacted about MGB research studies of whom 1,848 (14.5%) enrolled. “Physician invitations” were sent to 85,505 individuals of whom 5584 (6.5%) enrolled. “Volunteer invitations” were sent to 4,313 individuals (with or without prior connection to MGB), of which 2857 (64.2%) enrolled (Figure 3). The connection to the hospital system was critical in allowing us to invite enough individuals to achieve a fully enrolled, demographically representative cohort. This was particularly important for recruitment of Black and Hispanic/Latinx participants where regardless of invitation method, the enrollment rate was uniformly lower (Figure 3b). In total, 692 (75.0%) of Black and 579 (60.8%) of Hispanic/Latinx participants enrolled through “physician invitation.” Daily monitoring of enrollment trends was critical to guide community outreach and adjustments in distribution of invitations resulting in enrichment of under-represented demographic groups invited to join the study to achieve a representative cohort (Figure 4).

**Figure 3:**
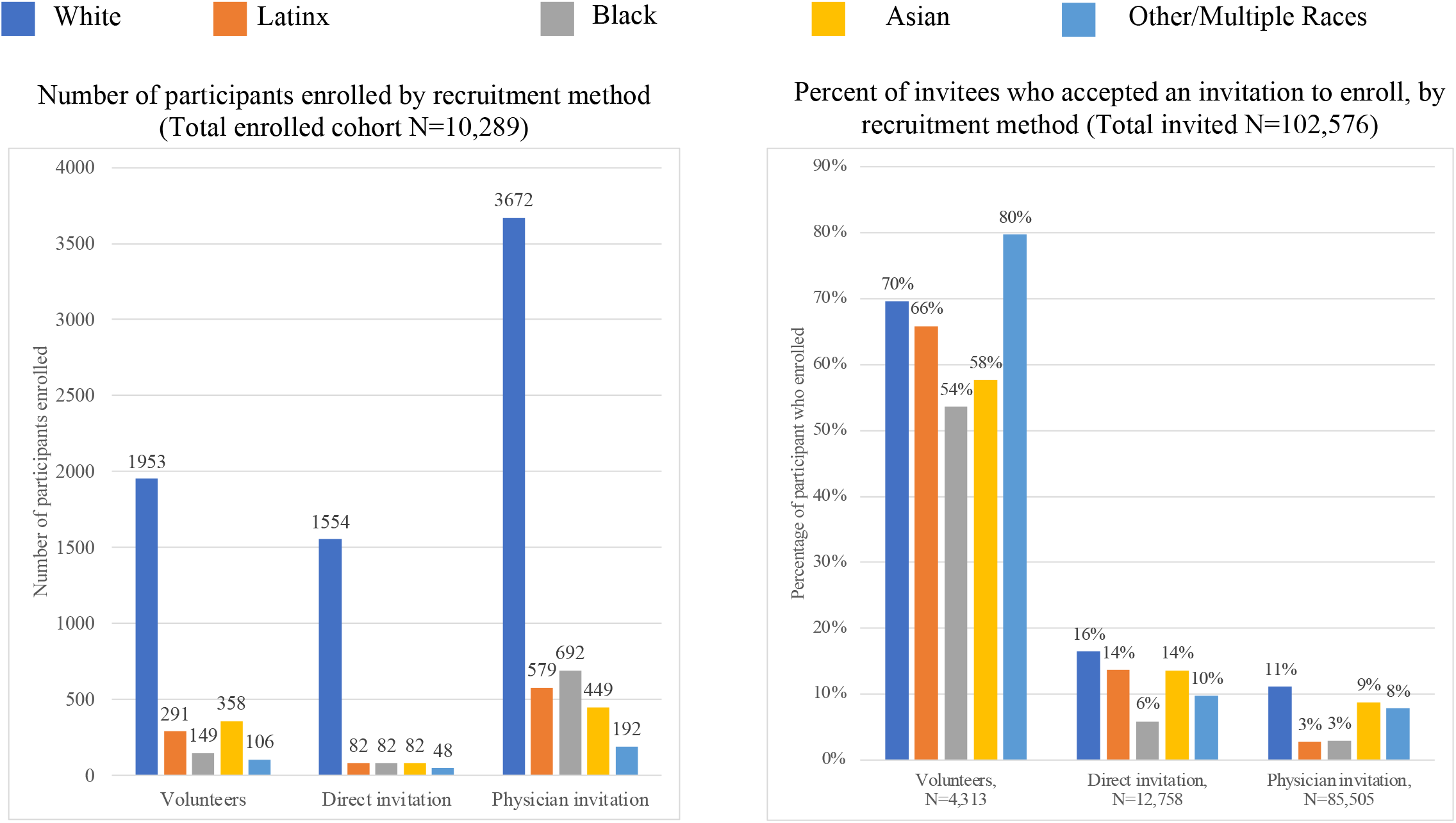
Enrollment by recruitment method.

**Figure 4:**
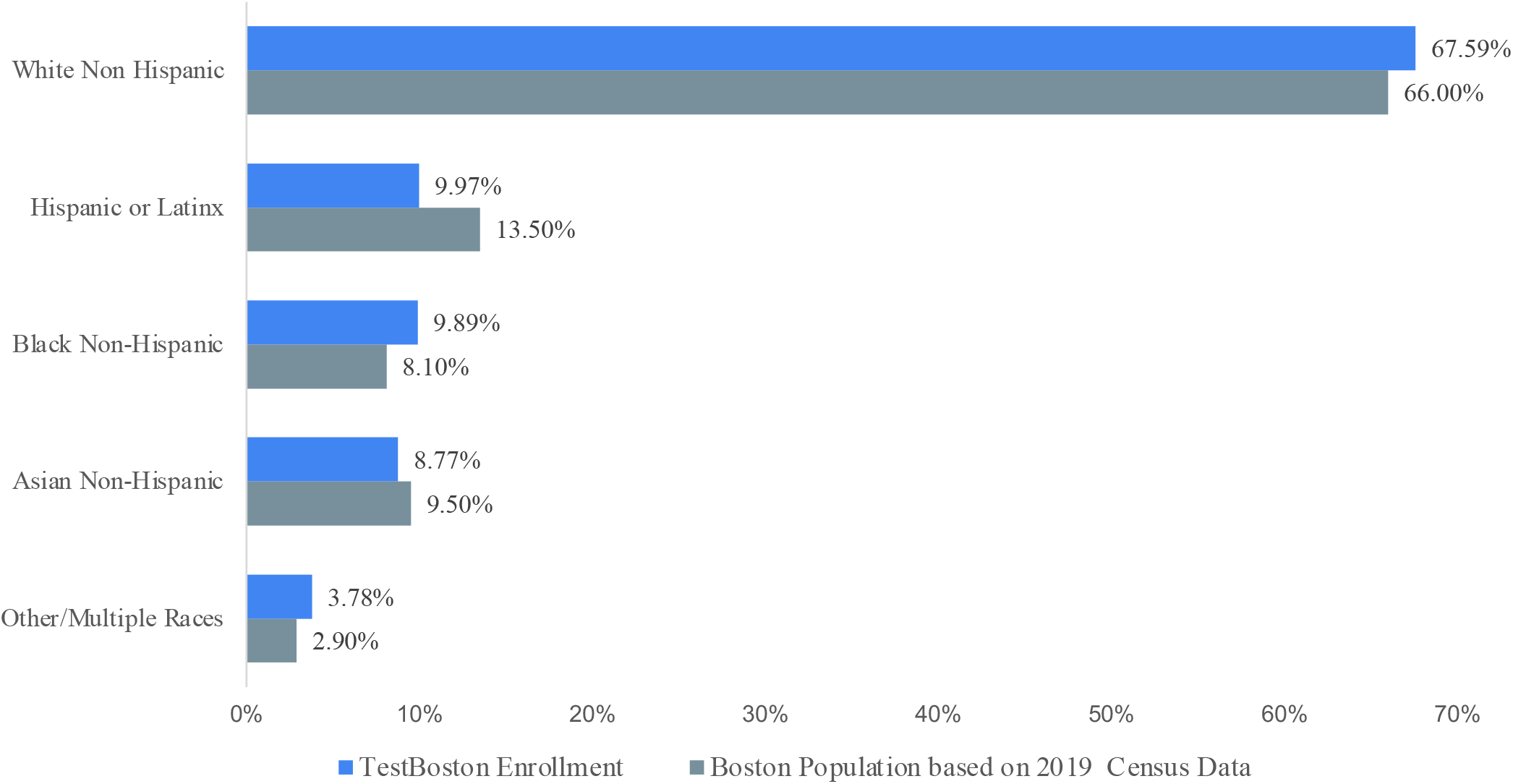
TestBoston cohort demographics compared to Boston census data.

We observed high levels of retention across demographic groups with some differences in numbers of kits returned as a measure of engagement and retention. TestBoston was designed for participants in the longitudinal cohort to return six monthly kits with an anticipated 10% attrition rate per month resulting in a final engagement rate of ∼50% after six months. Retention was slightly higher than expected with 5739 (55.8%) participants returning five or more kits within 30 days of receipt (Table 1). However, level of engagement and kit use behavior varied throughout the cohort and study period, mirroring the local COVID-19 surges and trends. We observed delays in participants completing kits due to travel, personal, or work obligations, as well as high volumes of kit use prior to holidays. The median number of days for return of monthly scheduled kits after delivery was six (IQR 2-13) compared to 3 days (IQR <1-8) for “on-demand” kits.

**Table 1:**
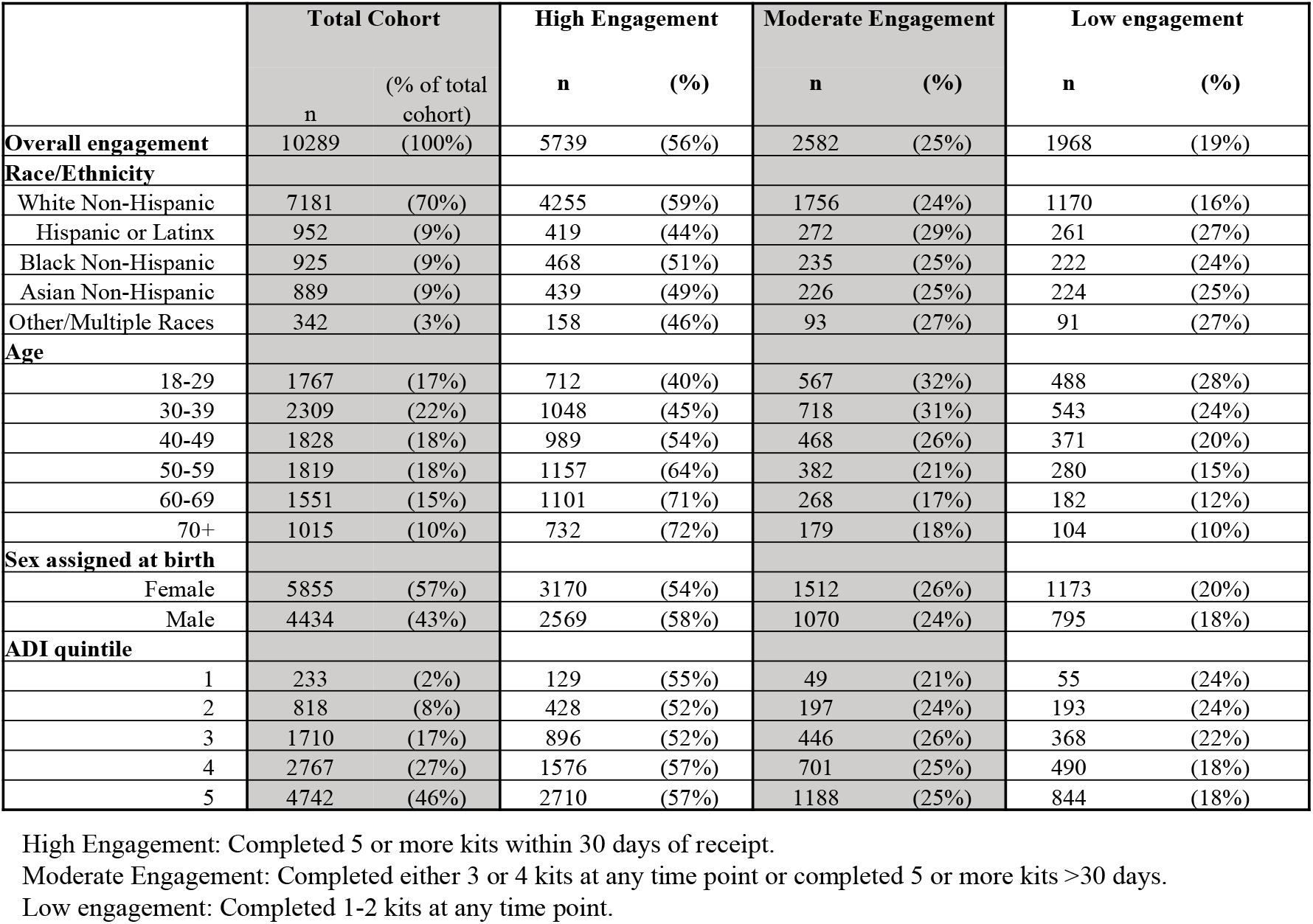
Longitudinal retention & engagement based on kit completion

In a multivariable model, there were modestly higher levels of engagement observed among participants who were White non-Hispanic (adjusted relative ratio [aRR], 1.11 compared with non-White or Hispanic; 95% CI 1.09 to 1.13), older (aRR, 1.06 for each 10 year increase; 95% CI 1.06 to 1.07), had a lower neighborhood disadvantage (aRR, 1.01 per ADI quintile increase; 95% CI 1.00 to 1.02), and those working remotely (aRR, 1.03 compared with unemployed or students; 95% CI 1.01 to 1.05). No statistically significant differences were observed by sex or those working in-person (Supplement 1).

### Study implementation exposed key areas of participant burden unique to remote trials models requiring intensive staff support

Participants successfully returned 44,277 test kits, of which 95.7% of nasal swabs were satisfactory and resulted. Compared to in-person trials where participants travel to study sites and are guided through procedures, TestBoston participants were required to independently navigate participation, including online registration, consent, survey completion, self-directed sample collection and shipment.

While the remote and automated nature of the study design reduced many tasks that would have been performed by study staff in traditional in-person visits, the additional burdens experienced by participants led to higher than anticipated study staff support requirements. Throughout the study, participants sent 11,500 emails. While the quantity of hotline calls and voicemails was not tracked, one to two full-time staff members were dedicated solely to answering hotline calls Monday through Friday during business hours. Trends in participant support needs changed over time (Figure 2c). Early inquiries included clarifying study aims, completing phone-assisted informed consent, and assisting participants without access to internet or with low digital and/or health literacy. As participants progressed through the study, requests became specific to kit completion such as forgotten passwords, kits lost in transit, and challenges in self-collection of samples. One of the most persistent support needs was assisting participants in completing their monthly surveys. Over 12,300 (28%) returned kits did not have a completed symptom survey at time of sample collection, necessitating study staff follow-up. Additional participant burdens included time required to complete the multi-step sample collection process, in particular the dried blood spot card, and difficulties adhering to the same-day sample shipping protocol required to ensure arrival at the lab within the requisite timeframe for accurate testing.

Intensive simultaneous support from six study team members was required to address high volume issues that arose, including uncontrollable, external events such as a worldwide Google cloud outage that impacted participant dashboard access, or nationwide holiday shipping delays that affected kit delivery, resulting in high volumes of participant inquiries. For participants with a positive COVID-19 diagnosis, physician support was critical in providing post-test counseling, assessing symptoms and risk profile, notifying the participant’s primary care physician, and referring for follow-up medical evaluation when needed. Responsibilities amongst dedicated team members were thus distributed based on type of support need such as software challenges, replacing lost or damaged kits, improving sample quality, and supporting the follow up of a positive COVID RT-PCR test (Figure 2d).

### A robust information technology infrastructure coupled with participant-informed data review enabled an adaptive, iterative approach to support engagement and data quality

Technological infrastructure and information technology (IT) automation were imperative for TestBoston implementation, scalability, monitoring of individual participant progress through study milestones, and tracking macro level trends in real-time. During the three-month enrollment period, the mean daily enrollment was 83 participants (range 2-914) and the mean daily test kits ordered was 330 (range 17-1684). At full enrollment, a daily mean of 525 completed kits (range 210-984), each individually barcoded and tracked, was returned to the testing lab. Pepper acted as a centralized clearing house of data to manage and track each step of participant progress starting with participant-entered survey data, to data from 3^rd^ party systems reporting kit order fulfillment, delivery status, processing and testing within the lab, and automated return of test results. This system was essential, for example, in rapidly identifying unused kits, allowing the study team to send reminder emails, offer collection support, or order replacements kits when needed. Throughout the study 1567 kits were replaced for those lost in transit, damaged, missing components, or otherwise misplaced.

The integration of the technology infrastructure, software engineers and study staff in the form of rapid communications, and daily reviews of study status and frequently asked questions enabled us to routinely monitor data and integrate feedback obtained through participant-staff support encounters. This led to near constant review and improvement, including both immediate resolution of challenges for individual participants and more systemic operational and software changes (Figure 2d). Standard operating procedures and template responses for over sixty frequently asked questions were modified in response to real-time data monitoring and participant challenges. Additionally, we were able to reduce the monthly unsatisfactory rates of returning nasal swabs for RT-PCR testing from 5.8% to 2.4% by instituting several processes including a dedicated study staff to review daily unsatisfactory results and contact participants to offer a re-test along with collection advice, changes to the kit design to simplify collection instructions, and changes to the shipping and receiving infrastructure.

## DISCUSSION

Recruitment, accrual, and retention are known challenges in biomedical research. We enrolled a cohort of over 10,000 individuals closely matching demographics of the greater Boston area with high levels of retention. Our model highlighted the critical role engaging physicians and leveraging connections to the medical system play in large-scale research recruitment in general, but specifically to ensure equitable representation. While the acceptance of an invitation was lower than among volunteers, the physician connection was critical in reaching demographic targets since the number of invitations was 12 times higher. However, disparities we observed in enrollment rates also highlight ongoing barriers not alleviated by the convenience of remote models, such as fear or research mistrust, and underscores the need for additional work to achieve equitable and representative enrollment.

The relatively lower retention rates observed among participants who were Black, Hispanic/Latinx, Asian, younger, and of higher neighborhood vulnerability suggest barriers once enrolled in remote trials may also be experienced disproportionally across demographic groups. While remote trials may alleviate structural barriers by allowing flexibility based on personal availability, TestBoston participants highlighted work and family obligations, time, and scheduling constraints that conflicted with sample return, as significant barriers. Other potential gaps that have been described include technology access, literacy and privacy concerns that may be unaddressed, and at worst, exacerbated by remote models (14,39–42). We attempted to mitigate these factors by providing translation services, intense participant support, and participant-informed adaptations. However, additional research is needed to further understand these barriers and identify solutions, along with a commitment to a participant-partnered approach (14,15,20,21).The TestBoston model enabled fast, efficient enrollment and collection of longitudinal infectious disease surveillance data at the height of the pandemic. This would not have been feasible using an in-person approach, which would have required over 44,000 discrete study visits. The approach decreased certain burdens for staff including scheduling visits, collecting and processing samples, and for participants including travel time and costs, that occur with traditional in-person visits. However, with the onus on participants to independently adhere to study protocols, a robust level of remote support was critical to optimize the participant experience and ensure sample and data quality. These per-participant “transferred burdens” (20) experienced by study staff lessened over time and as the cohort grew, ultimately resulting in a highly efficient model at scale.

Given the crucial role of technology and IT systems to enable a high volume of participants to self-report data and study staff to manage data, monitor participant progress, track kits as they pass through chain of custody, and respond to test results, the optimization of these systems in response to challenges in participant and staff usage required significant investment throughout the study. The most critical implementation lesson was the need for an adaptive approach. This was necessary given the inability to fully a priori anticipate all real-world challenges encountered throughout a remote study during a pandemic. Unanticipated challenges arose due to scaling, uncontrollable third-party events, or because real-world participant behavior deviated unpredictably from ideal behavior. While impossible to prepare for every contingency, it was critical to adopt a strategy to rapidly receive feedback and adapt.

## Limitations

Translation of our methods to other studies must be considered within the context of the unprecedented circumstances of the COVID-19 pandemic. Enrollment was likely enhanced by the attraction of receiving at-home COVID-19 testing at a time when Boston was entering a second surge of infections and access to testing was still limited. COVID-19 created an environment that was paradoxically both amenable to certain innovations and resistant to typically straightforward operations. TestBoston benefited from high levels of political will and a shared urgency from institutional and community stakeholders which helped to overcome barriers and delays in implementing a fully remote study.

While TestBoston succeeded in enrolling a demographically representative cohort based on Boston census data, there are shortcomings in using population as a benchmark for successful representation where a disease disproportionately impacts specific populations, in this case, Black and Hispanic/Latinx communities (43). There was inherent tension in trying to study the disease in an unbiased way and, concurrently wanting to help communities most affected by COVID-19 by providing greater access to testing. Finally, though our approach achieved engagement of those traditionally neglected from research, including those unable or unwilling to travel to in-person study visits, most participants were affiliated with the hospital network, and may not equally represent those who do not have access to healthcare.

## CONCLUSIONS

As shown by the TestBoston model, the decentralization of clinical trials offers tremendous potential to disrupt the clinical trial landscape by reaching more representative cohorts and increasing scale, reducing per-participant time commitments for study staff, and promoting accessibility. Such studies must appreciate the different set of challenges created, compared to in-person studies; as the responsibility to complete study activities is transferred to enrolled individuals, sufficient investment in resources in the form of participant support and software infrastructure are needed to ease participant burdens. These unique challenges will likely be universal across remote trial study design regardless of research area. Researchers undertaking remote models must prioritize continuous learning from participants at all stages, observing real-world experiences to ensure this potentially paradigm-shifting model does not create new, different barriers to inclusion, but rather is a true opportunity for more representative research involvement.

## Data Availability

All data produced in the present study are available upon reasonable request to the authors

## Acknowledgments

First and foremost, we would like to acknowledge our study participants from the greater Boston community, without whom this research would not have been possible. The authors would like to acknowledge the role of Mascon Medical (Woburn, MA), GBF (High Point NC), and UPS for their ongoing support for logistics and operations of this study. We further acknowledge the community leaders and physicians who supported recruitment and enrollment. We thank graphic designers Rahel Wachs and Anna Hupalowska, PhD for their contributions to graphic design and participant facing materials, and software engineers Yufeng Wang, Marco Ocana, Pegah Taheri, and Simone Maiwald.

---

**Supplement 1:**
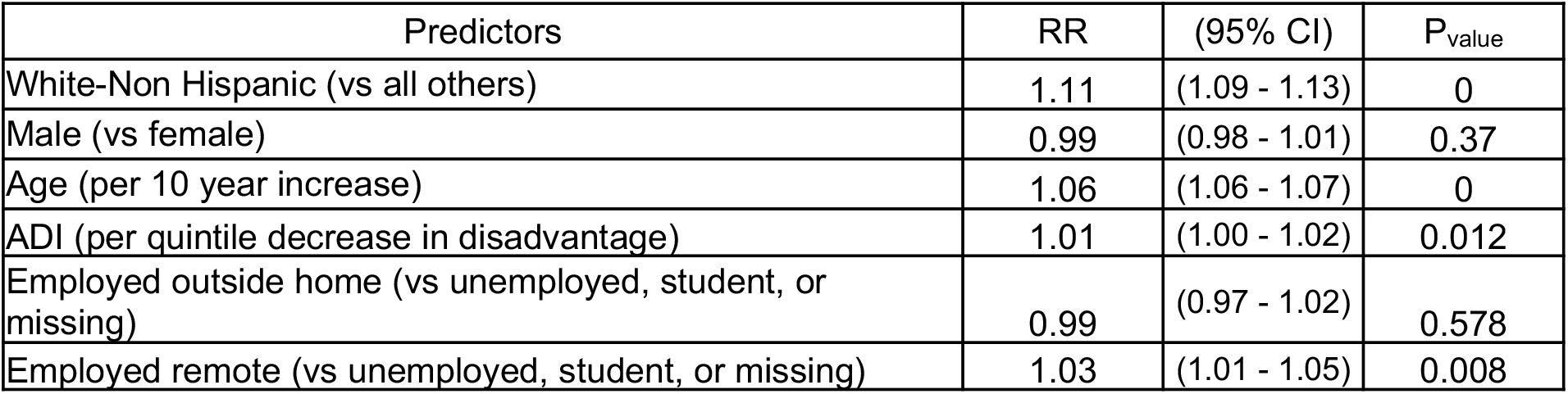
Longitudinal retention & engagement based on kit completion

| Predictors | RR | (95% CI) | P<sub>value</sub> |
| --- | --- | --- | --- |
| White-Non Hispanic (vs all others) | 1.11 | (1.09 - 1.13) | 0 |
| Male (vs female) | 0.99 | (0.98 - 1.01) | 0.37 |
| Age (per 10 year increase) | 1.06 | (1.06 - 1.07) | 0 |
| ADI (per quintile decrease in disadvantage) | 1.01 | (1.00 - 1.02) | 0.012 |
| Employed outside home (vs unemployed, student, or missing) | 0.99 | (0.97 - 1.02) | 0.578 |
| Employed remote (vs unemployed, student, or missing) | 1.03 | (1.01 - 1.05) | 0.008 |

